# Hemagglutination inhibition and alternate serologic responses following Influenza A(H3N2) virus infection

**DOI:** 10.64898/2026.04.21.26351404

**Authors:** Boshu Chen, Jose Victor Zambrana, Abigail Shotwell, Nery Sanchez, Miguel Plazaola, Sergio Ojeda, Roger Lopez, Daniel Stadlbauer, Guillermina Kuan, Angel Balmaseda, Florian Krammer, Aubree Gordon

**Author notes:** Corresponding authors: Florian Krammer & Aubree Gordon. These authors contributed equally (Boshu Chen, Jose Victor Zambrana). Co-senior authors (Florian Krammer, Aubree Gordon).

## Abstract

Background

Although the hemagglutination inhibition (HAI) titer remains the gold standard correlate of protection against influenza, it does not fully capture the broader antibody responses that contribute to immunity.

**Methods:** We analyzed immune responses in paired pre-infection and convalescent sera from 306 RT-PCR–confirmed A/H3N2 infections from two household studies (2014–18) in Managua, Nicaragua. Antibody responses were measured by HAI and enzyme-linked immunosorbent assays (ELISAs) against full-length hemagglutinin (HA), the HA stalk, and neuraminidase (NA). Participants were classified as HAI responders (≥4-fold HAI rise), alternate responders (no HAI rise but ≥4-fold boost in ≥1 ELISA), or no-response individuals (no ≥4-fold rise in any assay). We compared demographic, clinical, and pre-infection antibody characteristics across these groups. We also analyzed predictors of an NA response.

**Results:** Overall, 77% of participants had HAI seroconversion or a 4-fold rise. Among the 23% HAI non-responders, 62% had alternate antibody responses. No-response individuals had the highest pre-infection HAI and full-length HA titers (p < 0.0001), the lowest viral loads, and the fewest fever or influenza like illness (ILI) symptoms (p < 0.01). An NA response was more common among symptomatic individuals (p = 0.0483) and those with low or high baseline NA titers.

**Conclusions:** High baseline HAI titers can limit detectable 4-fold rises and are associated with milder illness. Evaluating additional immune responses may capture a more complete picture of the host response to infection, thereby improving surveillance and informing vaccine development.

## Introduction

Influenza virus continues to pose a significant global health burden, causing substantial morbidity and mortality through annual seasonal epidemics.^1^ Serologic studies are critical for measuring immunity and infection, and the hemagglutination inhibition (HAI) titer has long served as the gold standard for measuring antibody responses to influenza virus infection and vaccination. A ≥4-fold rise in HAI titer is typically used as serological confirmation of infection.^2,3^ In addition, HAI titers remain the only accepted correlate of protection for current influenza vaccines, with higher titers associated with reduced risk of infection and disease.^4–7^

The HAI assay detects antibodies directed against the variable hemagglutinin (HA) head domain and therefore provides a limited view of the antibody responses to infection.^8^ This limitation is particularly relevant for influenza A/H3N2 viruses, which undergo rapid antigenic drift and may present unique challenges for traditional serological assessment.^9^ Alternative antibody responses targeting the full-length HA protein, the conserved HA stalk domain, and neuraminidase (NA) represent potential targets for broadly protective vaccine development.^3^

Understanding the frequency and characteristics of individuals with HAI and alternate antibody responses is essential for understanding influenza virus immunity, protection, and improving surveillance accuracy.^10^ Here, we use data from two community-based influenza studies to characterize antibody responses to multiple influenza A (H3N2) virus antigens following infection. We assessed HAI antibody responses, full-length HA antibodies, HA stalk antibodies, and NA antibodies among RT-PCR–confirmed A/H3N2 infected individuals and identified immunologic and clinical factors distinguishing individuals who did or did not meet HAI-based infection detection criteria.

## Materials and Methods

### Study design

This study uses data from two household influenza studies conducted in Managua, Nicaragua. The Household Influenza Transmission Study (HITS) was a case-ascertained study conducted from 2012 to 2017.^11^ The Household Influenza Cohort Study (HICS) is an ongoing prospective family cohort study that began in 2017.^12^ In both studies, once influenza cases were confirmed by RT-PCR, their household members were monitored intensively for 10-14 days. Nasal and oropharyngeal swabs were collected on day 1 and then every 2–3 days throughout the monitoring period. Blood samples were collected both at the start of the monitoring period and again 30–45 days later. During the intensive monitoring period, daily symptom diaries were also collected.^11,12^ We analyzed RT-PCR-confirmed influenza A/H3N2 infections from the 2014, 2016 and 2017 influenza seasons.

### Laboratory methods

Swabs were tested by real-time RT-PCR following the validated Centers for Disease Control and Prevention protocols.^13^ Positive samples were also subtyped by RT-PCR. Paired pre- and post-exposure samples were assessed by using the hemagglutination inhibition (HAI) assay to determine antibody titers against A/Hong Kong/4801/2014.^14^ In addition, enzyme-linked immunosorbent assays (ELISAs) were used to quantify IgG antibodies targeting multiple antigens: full-length trimeric hemagglutinin (HA) from A/Hong Kong/4801/2014 (HK14), the HA stalk domain using a chimeric HA (cH7/3) construct with an H3 stalk (from HK14) and an H7 head (from A/Anhui/1/2013), and neuraminidase (NA) from HK14 in its tetrameric form.^15^ ELISAs were performed as described elsewhere.^16^ These antibody responses measured by ELISA are collectively referred to as “alternate antibodies” throughout the manuscript. Notably, A/Hong Kong/4801/2014 was the WHO-recommended H3N2 vaccine reference strain during the study seasons and provided standardized reagents for HAI and ELISA.^17^ Importantly, in this same Nicaraguan household cohort, antibody responses measured using the HK14 antigens have previously been identified as correlates of protection.^18^

### Statistical Methods

Participants were classified based on HAI response and the ratio of the pre- and post-exposure antibodies for full-length HA, HA stalk and NA. “HAI Responders” were defined as individuals with a ≥ 4-fold rise in HAI titer, while “HAI non-responders” were those who did not mount this level of response. HAI non-responders were stratified for further analysis as “Alternate responders”, defined as “HAI non-responders” who had a ≥ 4-fold response to full-length HA, HA stalk and/or NA, and “No response” individuals who did not exhibit a 4-fold rise to any of the serological tests.

We used chi-square tests, t-tests and analysis of variance (ANOVA) to compare group demographic and clinical characteristics. Clinical characteristics were defined as follows: fever was a measured temperature of ≥37.5 °C; influenza-like illness (ILI) required fever accompanied by either cough or sore throat; and acute respiratory infection (ARI) was defined as fever or any respiratory symptom. A locally estimated scatterplot smoothing (LOESS) regression with confidence intervals was used to visualize the relationship between HAI fold-changes and age. To quantify the odds of a ≥4-fold rise in NA, we fit a logistic regression model, adjusting for age, sex, clinical characteristics and pre-exposure NA levels with confidence intervals calculated via the Wald method. All hypothesis tests were two-sided and used an alpha level of 0.05. All statistical analyses and graphics were conducted using R 4.3.3 with the tidyverse package.^19,20^

## Results

### Study population

A total of 329 RT-PCR–confirmed influenza A/H3N2 virus infections were identified among 899 individuals enrolled from 169 A/H3N2-exposed households across the 2014, 2016, and 2017 seasons (Figure S1). Of these, 23 were excluded due to missing pre-and/or post-exposure HAI titer data because the blood sample was either not collected or insufficient volume was available for testing. The final analytic sample included 306 participants, of whom 207 (68%) were children (<15 years) and 99 (32%) were adults (≥15 years) (Figure S1, Table 1). Among them, 239 (78%) presented ILI, while 67 (22%) did not meet the ILI criteria (Table 1).

**Table 1.** HAI Responder versus Non-Responder characteristics.

HAI Responder versus Non-Responder characteristics.
| variable | Overall | HAI<br>Responder | HAI Non-Responder | P-value |
| --- | --- | --- | --- | --- |
| n | 306 | 235 | 71 |  |
| Male - n (%) | 138 (45.1%) | 103 (43.83%) | 35 (49.30%) | 0.500† |
| Age groups - n (%) |  |  |  | 1.00† |
| 0-14 years | 207 (67.6%) | 159 (67.66%) | 48 (67.61%) |  |
| 15+ years | 99 (32.4%) | 76 (32.34%) | 23 (32.39%) |  |
| Symptoms - n (%) |  |  |  |  |
| Fever | 245 (80.1%) | 195 (82.98%) | 50 (70.42%) | 0.0315† |
| ILI | 239 (78.1%) | 191 (81.28%) | 48 (67.61%) | 0.0228† |
| ARI | 287 (93.8%) | 223 (94.89%) | 64 (90.14%) | 0.241† |
| Cough duration (days) | 4.45 (6.87) | 4.76 (7.35) | 3.81 (4.94) | 0.210‡ |
| Pre-exposure antibody<br>levels - mean (SD) |  |  |  |  |
| HAI | 87.58 (195.84) | 56.3 (74.19) | 191.13 (366.82) | 0.00295‡ |
| Full-length HA | 331.75 (569.23) | 258.45<br>(353.51) | 574.38 (957.23) | 0.00797‡ |
| HA stalk | 48.98 (68.82) | 45.09 (67.84) | 61.86 (70.93) | 0.0806‡ |
| NA | 115.46 (289.69) | 93.86 (199.64) | 186.97 (475.04) | 0.112‡ |
| Cycle threshold - RT- | 25.4 (4.94) | 25.08 (4.76) | 26.46 (5.37) | 0.0594‡ |
| PCR - mean (SD) |  |  |  |  |
| †: Pearson's chi-square test; ‡: t-test, SD: Standard deviation |  |  |  |  |

Across all RT-PCR-confirmed influenza A/H3N2 virus infections (n=306), 77% exhibited a ≥4-fold rise in HAI titer, 83% in full-length HA, 54% in HA stalk, and 75% in neuraminidase (NA) antibodies (Table S1). Notably, 130 (42%) mounted a response for all four assays, 87 (28%) for three assays, 42 (14%) for two assays, 20 (7%) for one assay, and 27 (9%) had no response (Figure 1, Table S2).

**Figure 1.**
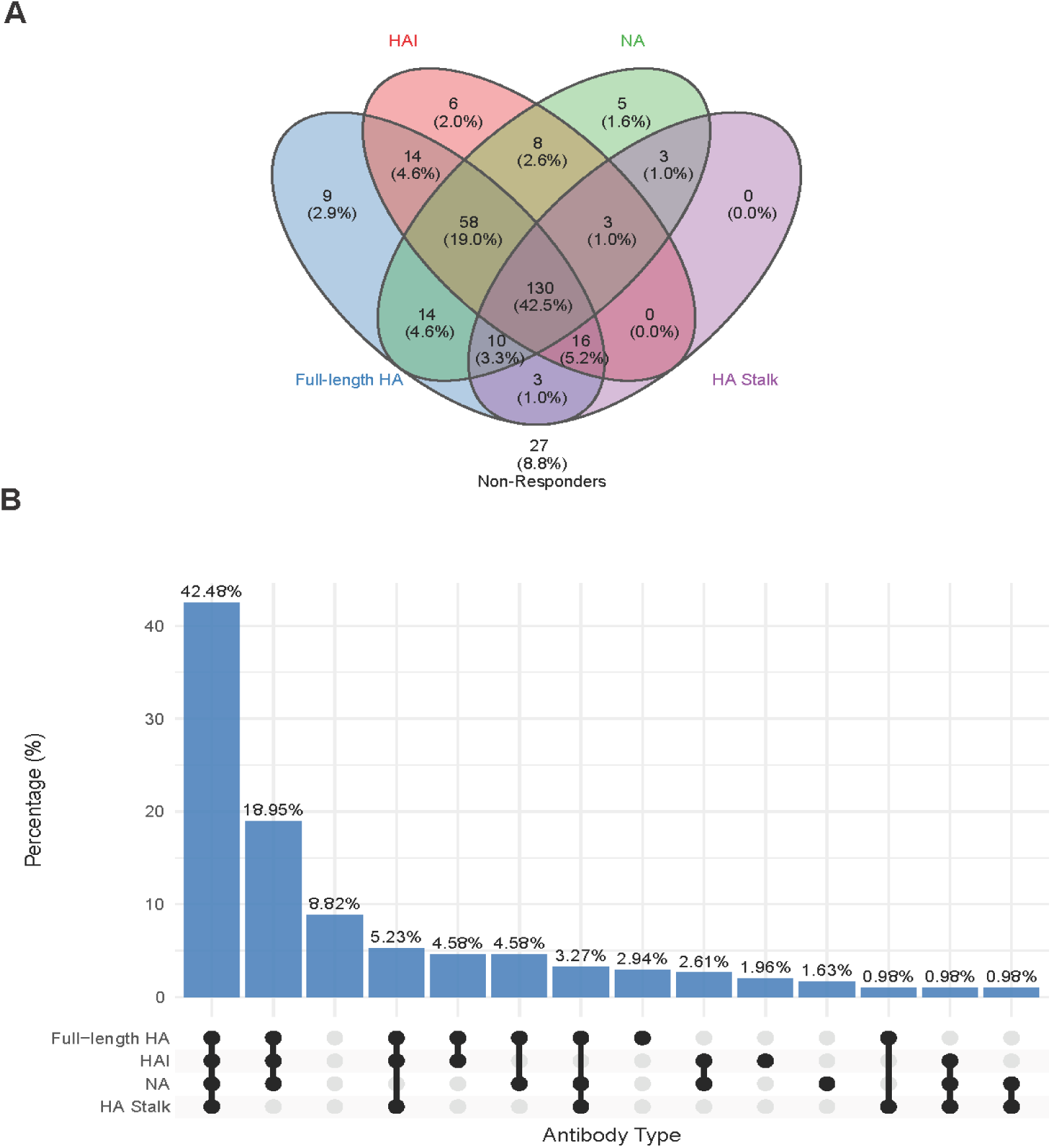
Distribution and overlap of ≥4-fold antibody responses by assay among A/H3N2-infected participants. (**A**) Venn diagram showing the overlap of ≥4-fold antibody rises across four assay types among the 306 A/H3N2-infected participants. Numbers represent the count (and percentage) of participants exhibiting a ≥4-fold rise in each individual assay or combination of assays. (**B**) UpSet plot illustrating the percentage of all observed combinations of ≥4-fold antibody responses across the four assay types.

### Comparison of HAI Responders and Non-Responders

Of the 306 participants, 235 individuals (77%) exhibited a ≥ 4-fold rise in HAI titer and were classified as “HAI responders” (Figure S1, Table 1). Among these 235 individuals, 103 (44%) were male, 159 (68%) were children (defined as ≤ 14 years of age), and 191(81%) met the symptom definition for ILI (Table 1). Additionally, 149 (63%) exhibited a ≥4-fold rise in anti-HA stalk antibodies, 218 (93%) in anti-full-length HA antibodies, and 199 (85%) in anti-NA antibodies (Table S1, Figure 1).

The remaining 71 (23%) individuals, who did not seroconvert in HAI titers, were defined as “HAI non-responders” (Table 1). Of these, 44 (62%) individuals demonstrated a ≥ 4-fold rise in antibodies against HA stalk, full-length HA and/or NA and were further classified as “alternate responders”, while 27 (38%) individuals, who showed no ≥4-fold rise against any of the antibody measures, were categorized as “no response” (Table 2). Among these alternate responders, 21 (47%) were male, 30 (68%) were children and 33 (75%) had ILI (Table 2). Among the no response group, 14 (52%) were male, 18 (67%) were children and 15 (56%) had ILI (Table 2). There were no statistically significant differences in age (p = 1.00) or sex (p = 0.500) between antibody responders and non-responders (Table 1). However, HAI non-responders were significantly less likely to report fever (p = 0.031) or to meet the case definition for ILI (p = 0.023), suggesting a milder clinical presentation (Table 1). Pre-exposure antibody titers against HAI and alternate responses were significantly higher in HAI non-responders compared to HAI responders (HAI, p = 0.00295; full-length HA, p = 0.00797; HA stalk, p = 0.0806; and NA, p = 0.112) (Table 1, Figure 2). Additionally, across all age groups, HAI non-responders exhibited lower fold-change titers against full-length HA, HA stalk and NA compared to HAI responders (Figure S2).

**Figure 2.**
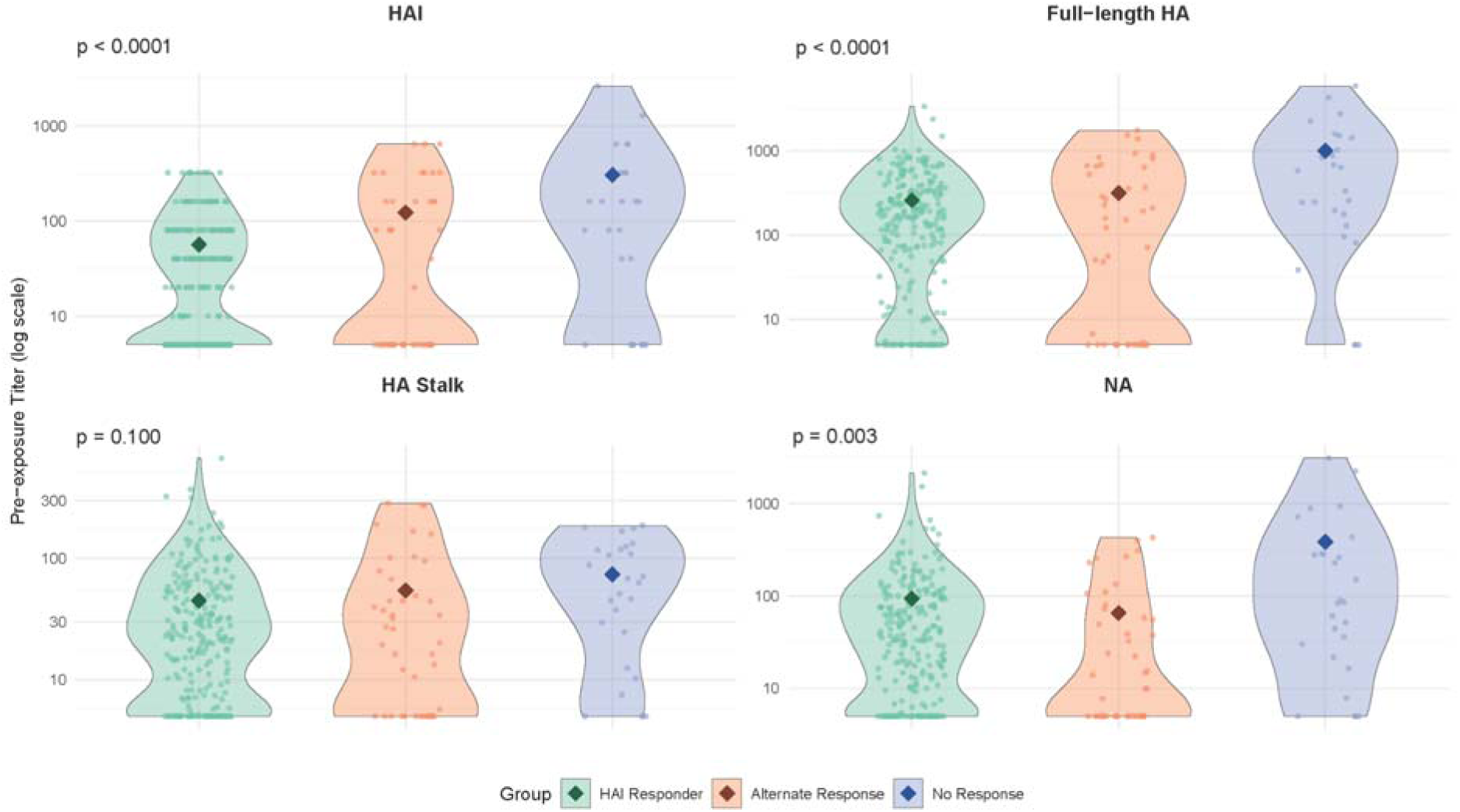
Pre-exposure antibody titers by assay among HAI responders, alternate responders, and no response individuals. Violin plots show the distribution of pre-exposure antibody titers (log scale) across three participant groups: HAI responders, alternate responders, no response individuals. Each panel represents a different assay. Individual data points and group means (diamonds) are displayed within each violin. P-values correspond to global comparisons across groups using one-way ANOVA.

**Table 2.**
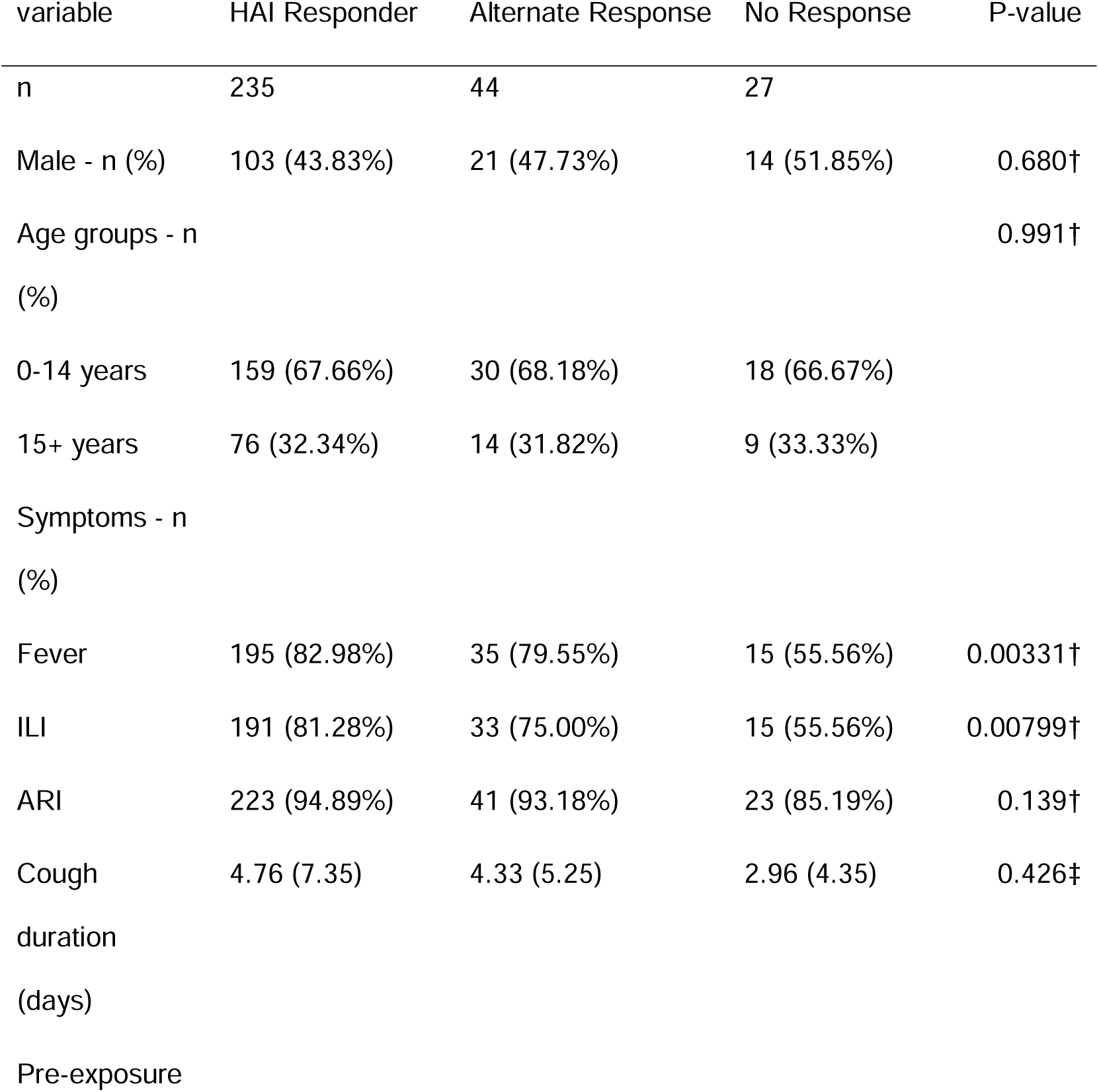
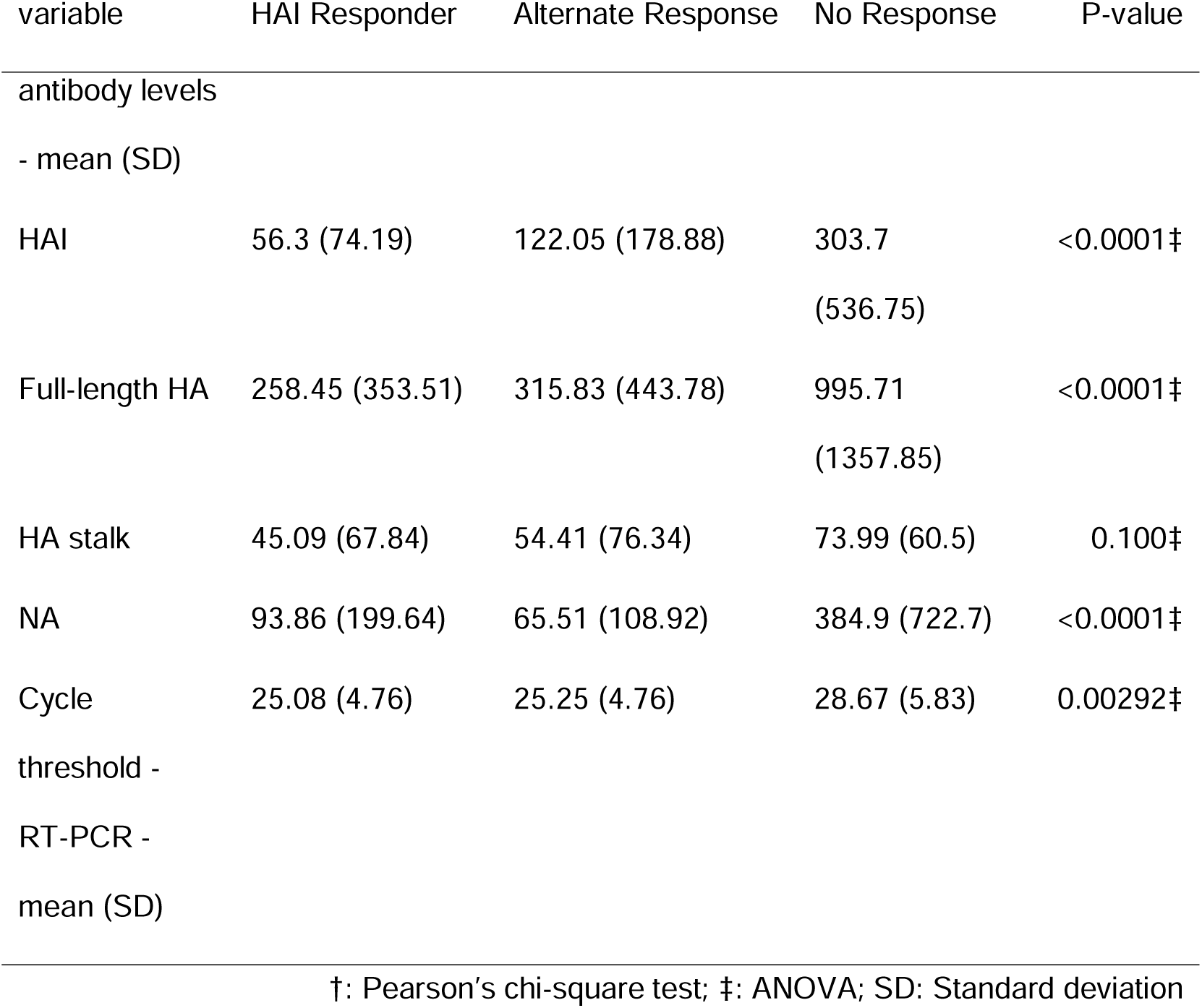
HAI Responders versus Alternate responders versus No response.

### No response individuals compared to HAI responders and alternate responders

We further split HAI non-responders into two groups: those who had an alternate antibody response (alternate responders; n=44) and those who did not exhibit a 4-fold rise in any antibody level (no response, n=27) (Figure S1, Table 2). Overall, there were no significant differences in age or sex across groups (Table 2). However, individuals in the no response group were less symptomatic than both HAI responders and alternate responders, with only 56% reporting fever (p = 0.00331) and meeting the symptom profile for ILI (p = 0.00799) (Table 2). Differences in pre-exposure antibody titers were observed across groups. No response individuals (mean HAI titer: 303.7) and alternate responders (mean HAI titer: 122.05) had higher pre-exposure HAI titers compared to HAI responders (mean HAI titer: 56.3; p < 0.0001) (Table 2, Figure 2). Similarly, pre-exposure antibody levels against full-length HA (p < 0.0001) and NA (p < 0.0001) were significantly higher in the no response group compared to HAI responders and alternate responders (Table 2, Figure 2).

When comparing alternate responders to HAI responders, pre-exposure HAI titers were significantly higher in alternate responders (p = 0.0001) (Table S3). There was also a trend towards higher pre-exposure antibody levels against full-length HA and HA stalk, and lower levels against NA among alternate responders, though these differences were not statistically significant (Table S3). Symptom prevalence did not differ significantly between alternate responders and HAI responders. The majority (97%) of HAI responders showed a ≥ 4-fold rise in at least one alternate antibody, with a higher frequency of ≥4-fold responses to any alternate antibody compared to alternate responders (Figure 1, Table S1). When examining the minimum RT-PCR cycle threshold (Ct) value during the infection, the no-response group (Mean = 28.67) showed a significantly (p = 0.00292) higher mean Ct than the HAI responders (Mean = 25.08) and alternate responders (Mean = 25.25), indicating that this group carried the lowest viral load (Table 2).

### Neuraminidase and symptom characteristics

Given growing interest in incorporating NA into conventional influenza vaccines and its status as a comparatively accessible target for improving breadth, we examined demographic and clinical factors associated with NA antibody responses.^21^ Age and sex were not significantly associated with an NA rise. A multivariable logistic regression analysis, adjusting for acute respiratory infection, age, sex and pre-exposure NA levels showed ARI was significantly associated with an NA rise (OR 2.85, p = 0.0483). Additionally, individuals with no or very low (≤5; OR 0.21, 95% CI 0.11–0.40, p = 0.00125) or high (≥81; OR 0.28, 95% CI 0.13–0.61, p < 0.0001) pre-exposure NA antibody levels had lower odds of experiencing a 4-fold rise compared with those with moderate baseline levels (6–80).

## Discussion

Our findings highlight that reliance on HAI alone provides an incomplete picture of humoral influenza virus immune responses against influenza A H3N2 virus infection. We found that nearly one-quarter of RT-PCR confirmed H3N2 infections did not elicit a 4-fold rise in HAI antibodies. These individuals were characterized by higher baseline antibody titers, milder symptoms, and lower viral loads. In particular, the no-response group, defined as individuals with <4-fold rises in HAI titers and any alternate antibody, had elevated pre-exposure HAI and full-length HA antibodies and was less likely to report fever or meet the ILI definition. Taken together, these findings suggest that reliance on HAI titer alone may underestimate H3N2 infections, particularly among individuals with high baseline antibody levels from exposures or with mild clinical presentations.

When comparing our H3N2 findings with prior H1N1 results from the same prospective studies analyzed using the same methods, we observed both similarities and differences.^10^ This parallel design makes it possible to directly contrast immune responses across influenza A virus subtypes, which is rarely feasible in other settings. In both studies, roughly one-quarter of infections failed to mount a 4-fold rise in HAI titers, and more than half of these non-responders demonstrated response to at least one alternate antibody. In both H1N1 and H3N2, the no response group was characterized by high baseline antibody titers and milder clinical illness, suggesting that individuals with strong pre-existing immunity experience attenuated infections without detectable serologic response.

In contrast, the profile of alternate responders diverged between studies. For H1N1, alternate responders had lower antibody levels than HAI responders, whereas for H3N2, alternate responders exhibited higher pre-exposure HAI titers and a greater proportion had detectable HAI antibodies at baseline. This pattern likely reflects greater antigenic drift of H3N2 viruses, where preexisting antibodies may be less protective and more frequently associated with non-response by HAI.^21^ It is also important to note that the H1N1 study was conducted in 2013 and 2015, shortly after the H1N1 pandemic, when most individuals would have encountered the pandemic H1N1 strain only once and carried preexisting seasonal H1N1 immunity. Thus, participants had likely experienced at most one pandemic H1N1 infection, a context drastically different from the repeated exposures characteristic of H3N2. Additionally, H1N1 viruses undergo slower antigenic change, and high baseline titers appear to be more reliably protective, reducing the likelihood that such individuals would become infected and appear as non-responders.^18^

Our observation that a substantial fraction of H3N2 infections did not produce a 4-fold rise in HAI titer is consistent with prior reports. Several cohort and challenge studies have documented that individuals with high baseline titers often do not demonstrate conventional serologic responses despite virologically confirmed infection.^22–24^ This raises important questions about the sensitivity of HAI assay for capturing immune boosting, particularly for H3N2, which is known for more antigenic.^9^ Recent studies have also emphasized the role of non-HAI antibodies, including those directed against full-length HA and neuraminidase, in shaping protection and influencing clinical outcomes.^8,10,21,25,26^ Our findings add to this literature by demonstrating, in a community-based prospective cohort and transmission study, that alternate antibody responses can be detected in a majority of individuals that do not seroconvert to HAI, reinforcing the value of using multiple immunologic markers.

Moreover, baseline NA antibody levels and developing ARI symptoms emerged as key determinants of neuraminidase (NA) boosting after an influenza A H3N2 virus infection. Individuals with high pre-existing NA titers showed attenuated boosts, consistent with a ceiling effect on antibody amplification.^27^ This consideration is especially relevant as neuraminidase is increasingly recognized for inclusion in broadly protective influenza vaccine formulations.

This study has several limitations. The sample size for some subgroups, particularly the no response group, was relatively small, which may have limited our statistical power to detect significant differences and associations. Additionally, there is potential for misclassification of very short infections that may not have been adequately captured during the intensive monitoring period, although individuals are sampled every two to three days. Furthermore, our study focused exclusively on systemic antibody responses and did not assess mucosal or cellular immunity.

Our findings show that a substantial proportion of RT-PCR-confirmed influenza A/H3N2 virus infections do not elicit the expected ≥4-fold HAI response, yet still display robust immune activation as assessed by alternative serological markers. Recognizing these alternate responders highlights the complexity of influenza virus immunity and suggests that current surveillance methods based solely on HAI titer may underestimate the infection rate and could bias results. Moreover, we observed that higher pre-infection antibody levels correlate with reduced HAI responses and milder clinical courses, highlighting how baseline immunity can shape both disease severity and subsequent serological profiles. Together, these findings argue for incorporating broader serological assessments in future influenza surveillance and research.

## Author contributions

BC and JVZ contributed to data curation, formal analysis, methodology, visualization, and writing of the original draft. AS contributed to data curation. NS, MP, SO, RL, and DS contributed to investigation. GK and AB contributed to project administration and investigation. FK and AG contributed to conceptualization, funding acquisition, methodology, and supervision. All authors contributed to writing—review and editing.

## Acknowledgements

This work was supported by the National Institute of Allergy and Infectious Diseases (R01 AI120997 to A.G.; contracts HHSN272201400008C and 75N93019C00051 to F.K., HHSN272201400006C and 75N93021C00016 to A.G.). Additionally, this work was supported, in part, by Flu Lab (award to A.G.). We are grateful to the dedicated study teams in Nicaragua at the Centro Nacional de Diagnóstico y Referencia and the Sócrates Flores Vivas Health Center. The funders had no role in the conduct or reporting of the study.

## Competing interests

The Icahn School of Medicine at Mount Sinai has filed patent applications relating to SARS-CoV-2 serological assays, NDV-based SARS-CoV-2 vaccines influenza virus vaccines and influenza virus therapeutics which list FK as co-inventor and FK has received royalty payments from some of these patents. Mount Sinai has spun out a company, Castlevax, to develop SARS-CoV-2 vaccines. FK is co-founder and scientific advisory board member of Castlevax. FK has consulted for Merck, GSK, Sanofi, Gritstone, Curevac, Seqirus and Pfizer and is currently consulting for 3rd Rock Ventures and Avimex. The Krammer laboratory is also collaborating with Dynavax on influenza vaccine development. DS is an employee and shareholder of Moderna. AG reports institutional research funding from Flu Lab and Open Philanthropy; personal honoraria from Hope College and the La Jolla Institute of Immunology; compensation for expert testimony from Berman and Simmons; and travel support from the Gates Foundation and the National Institutes of Health (NIH). AG has also served, or currently serves, in an advisory capacity for Janssen Pharmaceuticals and Sanofi Pasteur. All other authors declare no potential competing interests.

## Data availability

Researchers seeking access to the study data are encouraged to submit a formal request to A.G. or to the Committee for the Protection of Human Subjects at the University of Michigan. To ensure ethical oversight and appropriate data use, all requests will be reviewed and approved on a case-by-case basis. Because the data include information collected in Nicaragua, access is subject to Nicaraguan data ownership regulations and may require approval from the appropriate Nicaraguan authorities. Final decisions are expected within approximately two months. Requests for materials and related correspondence should be addressed to AG.

## Ethics approval and patient consent

This study was approved by the University of Michigan Health Sciences and Behavioral Sciences Institutional Review Board (HUM00091392, HUM00119145) and the Nicaraguan Ministry of Health Institutional Review Board. Informed consent was obtained for all participants and verbal assent was obtained from children aged ≥6 years.

