## Supplemental material for "Hemagglutination inhibition and alternate serologic responses following Influenza A(H3N2) virus infection"

These authors contributed equally.

### Supplemental Tables

**Table S1** - Proportion of 4-fold rises among HAI Responders vs Alternate Responders by alternate assays

| Variable | Overall | HAI Responder | Alternate Response | P-value |
| --- | --- | --- | --- | --- |
| n | 306 | 235 | 44 |  |
| Full-length HA - n (%) | 254 (83.01) | 218 (92.77) | 36 (81.82) | 0.0408 |
| HA stalk - n (%) | 165 (53.92) | 149 (63.40) | 16 (36.36) | 0.0015 |
| NA - n (%) | 231 (75.49) | 199 (84.68) | 32 (72.73) | 0.0872 |
| Any alternate - n (%) | 273 (89.22) | 229 (97.45) | 44 (100.00) |  |
| P-values calculated by Pearson’s chi-square test | | | | |

**Table S2** - Distribution of ≥4-fold antibody responses by number of overlapping assays

| **Number of responses** | **Count (percentage)** |
| --- | --- |
| 0 | 27 (8.82%) |
| 1 | 20 (6.54%) |
| 2 | 42 (13.73%) |
| 3 | 87 (28.43%) |
| 4 | 130 (42.48%) |
| **Total** | **306 (100%)** |

**Table S3** - HAI Responders versus Alternate responders

| Variable | HAI Responder | Alternate Response | P-value |
| --- | --- | --- | --- |
| **n** | 235 | 44 |  |
| **Male** | 103 (43.83%) | 21 (47.73%) | 0.7549† |
| **Age** |  |  | 1.0000† |
| 0-14 years | 159 (67.66%) | 30 (68.18%) |  |
| 15+ years | 76 (32.34%) | 14 (31.82%) |  |
| **Symptoms** |  |  |  |
| Fever | 195 (82.98%) | 35 (79.55%) | 0.7388† |
| ILI | 191 (81.28%) | 33 (75.00%) | 0.4508† |
| ARI | 223 (94.89%) | 41 (93.18%) | 0.9220† |
| Cough duration (days) | 4.76 (7.35) | 4.33 (5.25) | 0.7085‡ |
| **Pre-exposure antibody levels** |  |  |  |
| HAI | 56.3 (74.19) | 122.05 (178.88) | 0.0001‡ |
| Full-length HA | 258.45 (353.51) | 315.83 (443.78) | 0.3446‡ |
| HA stalk | 45.09 (67.84) | 54.41 (76.34) | 0.4128‡ |
| NA | 93.86 (199.64) | 65.51 (108.92) | 0.3605‡ |
| †: Pearson’s chi-square test; ‡: t-test | | | |

**Table S4** - Serological and Symptom Characteristics, Along With the Odds of a ≥4-Fold NA Rise

| Variable | Total (Column%) | >=4 fold (Row%) | p-value† | OR estimate (95% CI)‡ | p-value‡ |
| --- | --- | --- | --- | --- | --- |
| **n** | 306 (100.00%) | 231 (75.49%) |  | - | - |
| **Fever** |  |  | 0.396 |  |  |
| Yes | 245 (80.07%) | 188 (76.73%) |  | - | - |
| No | 61 (19.93%) | 43 (70.49%) |  | - | - |
| **ILI** |  |  | 0.323 |  |  |
| Yes | 239 (78.10%) | 184 (76.99%) |  | - | - |
| No | 67 (21.90%) | 47 (70.15%) |  | - | - |
| **ARI** |  |  | 0.117 |  |  |
| Yes | 287 (93.79%) | 220 (76.66%) |  | 2.85 (1.01-8.07) | 0.0483 |
| No | 19 (6.21%) | 11 (57.89%) |  | ref |  |
| **Age (years)** |  |  | 0.432 |  |  |
| 0-14 | 207 (67.65%) | 153 (73.91%) |  | ref | 0.554 |
| 15+ | 99 (32.35%) | 78 (78.79%) |  | 1.21 (0.62-2.36) |  |
| **Male** | 138 (45.10%) | 104 (75.36%) | 1.000 | 0.91 (0.52-1.59) | 0.737 |
| **Pre-exposure NA titer** |  |  | 0.000 |  |  |
| 5 | 69 (22.55%) | 47 (68.12%) |  | 0.21 (0.11-0.40) | 0.00125 |
| 6-80 | 140 (45.75%) | 124 (88.57%) |  | ref |  |
| 81+ | 97 (31.70%) | 60 (61.86%) |  | 0.28 (0.13-0.61) | <0.0001 |
| †: Pearson’s chi-square test; ‡: Multivariate logistic regression | | | | | |

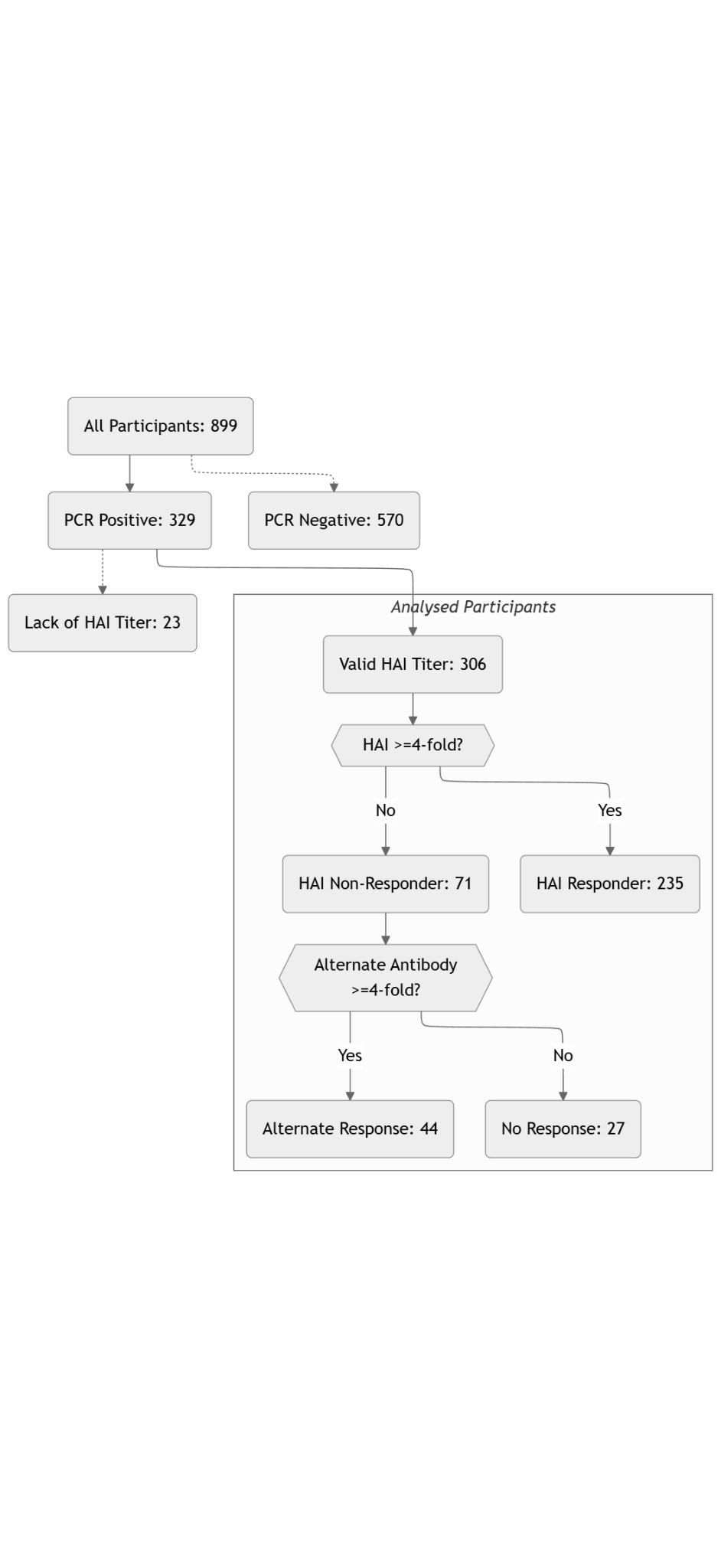

**Figure S1** - Participant selection and classification of HAI and alternate antibody responses.

Flow diagram summarizing participant inclusion and response classification.

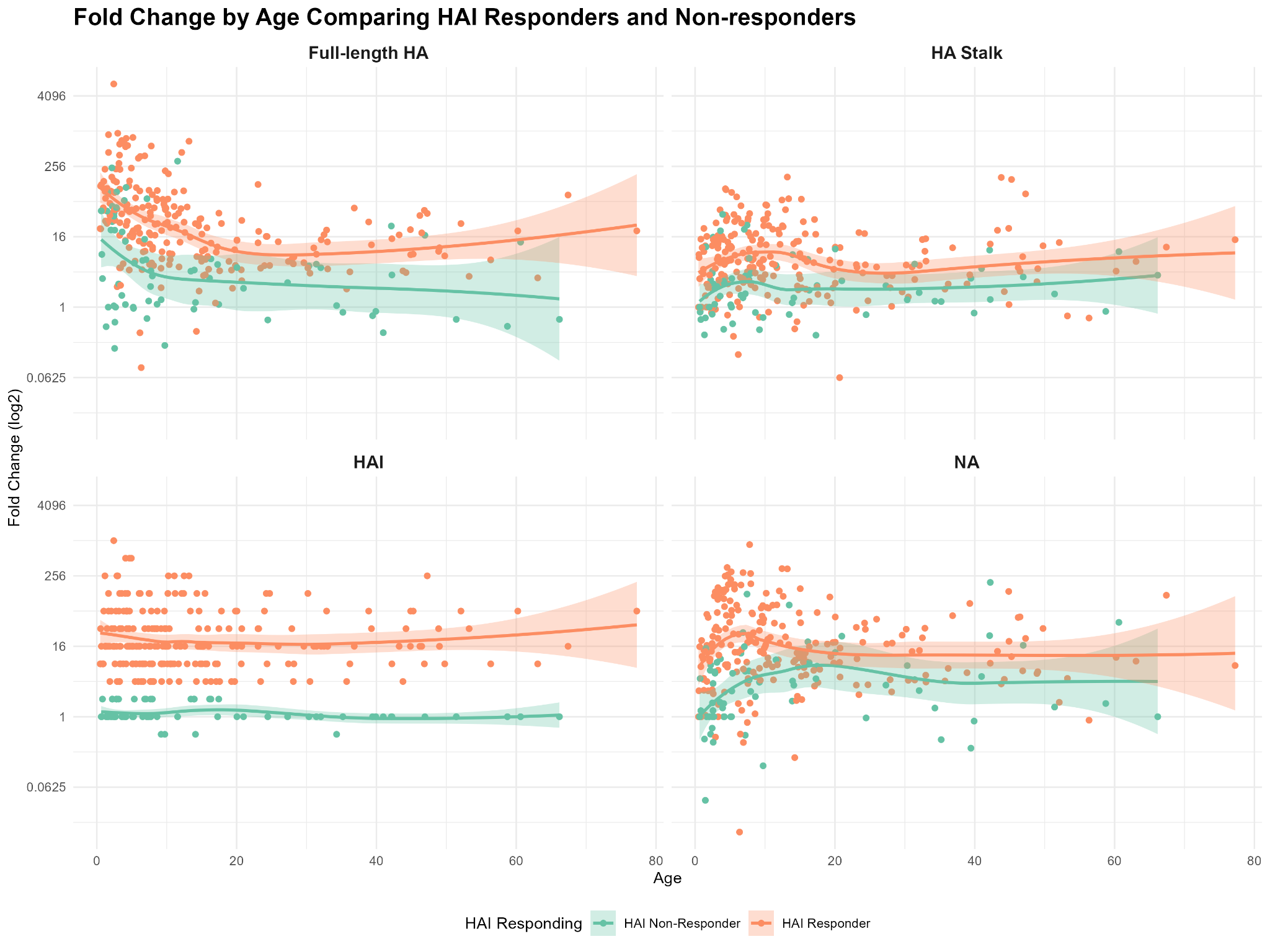

**Figure S2** *- Fold-change by age comparing HAI responders and Non-responders*

Scatterplots show the distribution of fold-change values across age for HAI responders and non-responders. Each panel represents a different assay. LOESS-smoothed curves with 95% confidence intervals illustrate trends by age in antibody fold-changes within each responder group.
